# A Cross-Sectional Survey of Australian Women’s Perspectives and Experiences of Exercise During Pregnancy, Including Women that Experience Mental Illness

**DOI:** 10.1101/2023.07.25.23292807

**Authors:** Noor S. Jarbou, Kimarnie Baskerville, Mariam Gabra, Tess Mawson, Kelly A. Newell, Jessica Nealon

**Author notes:** joint corresponding authors: Kelly A. Newell; Jessica Nealon.

## Abstract

**Purpose:** The aim of this study was to develop an understanding of Australian women’s perspectives and experiences around exercise during pregnancy, including women that experience mental illnesses such as depression.

**Method:** An anonymous online survey of women, administered via Qualtrics Australia, was open for 4 weeks between November - December 2021. The survey consisted of a 45-item questionnaire collecting data on demographics (including pregnancy status), attitudes/beliefs regarding conducting exercise during pregnancy, knowledge of exercise in pregnancy guidelines, exercise in pregnancy experience and mental health experiences of responders during pregnancy. Analysis of responses were restricted to those who have experienced a pregnancy. Descriptive statistics and frequency tables were calculated for all questions. Pearson’s Chi-Squared tests were used to determine the differences in response by mental health status (*p* < 0.05).

**Results:** There were 695 eligible responses. Most responders believe that regular exercise during pregnancy is safe for mother and baby (94%), would help prevent a decline in a pregnant person’s mood (88%) and help to improve a pregnant person’s mood (92%). The majority of responders (71%) were not aware of the WHO and Australian Government Department of Health recommendations for conducting exercise during pregnancy. 68% of responders conducted exercise during all or part of their most recent pregnancy. However, there was a significant association between a reported diagnosis of a mental illness during their lifetime and participation in structured exercise during pregnancy (*p*=0.009), with fewer individuals with a mental illness exercising during pregnancy compared to those without (61 vs 71%). Despite the majority of respondents participating in exercise during pregnancy, more than half of responders report conducting less exercise than that recommended by current guidelines. Fifty-seven percent of responders recall being advised to exercise during their most recent pregnancy, mostly by their GP (54%), midwife (46%) and obstetrician (38%). Walking was the most advised exercise type (80%) followed by swimming (46%). However, 69% of responders report receiving no or little advice from their healthcare provider about the safety and benefits of exercise during their most recent pregnancy, but 45% of responders felt satisfied about the advice they did receive.

**Conclusion:** This study has shown that Australian women believe that exercise during pregnancy is safe and has benefits for mental health. However, many women report receiving little advice about this from their healthcare provider. Furthermore, women report not being aware of or meeting the WHO/National guidelines regarding exercise in pregnancy. Women do report primarily conducting low intensity exercise during pregnancy (walking, swimming, yoga). Importantly, fewer respondents with a diagnosed mental illness report exercising during pregnancy. Considering the potential benefits of exercise for mental illness, employing strategies to increase engagement with exercise during pregnancy is important. Further research to determine strategies to increase exercise in pregnant women, particularly those with a mental illness, are needed.

## Introduction

Exercise during pregnancy has been shown to have numerous benefits for women including decreased risk of developing gestational diabetes (Abirami and Judie, 2014; de Barros et al., 2010; Koivusalo et al., 2015), hypertension and preeclampsia (Kasawara et al., 2012; Martin and Brunner Huber, 2010; Spracklen et al., 2016), reduced risk of excessive gestational weight gain (Bennett et al., 2018; Hinman et al., 2015; Hopkins and Cutfield, 2011), improved cardiovascular function (Witvrouwen et al., 2020), and prevention of or improvement in lower back and pelvic pain during pregnancy (Davenport et al., 2019b). Emerging evidence also suggests exercise in pregnancy is beneficial for mental health. In our recent systematic review we showed that structured exercise (including resistance training, walking, and aquatic exercise) during pregnancy in those with no diagnosed mental illness, reduced anxiety and depressive scores when measured during and after pregnancy (Jarbou and Newell, 2022). In the limited studies that have examined exercise in pregnancy in depression, the exercise interventions have been restricted to yoga, with the evidence regarding its ability to reduce anxiety and depressive-like symptoms inconclusive (Jarbou and Newell, 2022).

The World Health Organisation (WHO), Exercise and Sports Science Australia (ESSA) and the American College of Obstetrics and Gynaecology (ACOG) recommend that pregnant women, without contraindications(e.g. preeclampsia, premature labour, incompetent cervix), participate in at least 150 mins of moderate intensity exercise or 75 mins of vigorous intensity exercise per week (or an equivalent combination thereof), or exercise at least five days per week (e.g., completing 30-min of exercise for 5 days a week to perform 150 mins) (Bull et al., 2020). Women should avoid contact sports, sit-ups, risk of falls or supine exercises, but are encouraged to participate in Pilates or yoga, walking, stationary cycling, and aquatic based activities (Exercise and Sports Science Australia 2018, p. 25). These recommendations are also reflected in the Australian Department of Health guidelines for physical activity during pregnancy (Australian Goverment Department of Health, 2021).

Despite the potential benefits of exercise and the guidelines recommending exercise during pregnancy, pregnant women are not meeting the recommended guidelines for exercise worldwide (Gaston and Vamos, 2013; Gjestland et al., 2013; Hesketh and Evenson, 2016; Mottola et al., 2018). Recently, the Australian Institute of Health and Welfare (AIHW) reported that only 30% of pregnant women met the physical activity recommendations of WHO and the Australian Department of Health (Australian Institute of Health and Welfare, 2019). On the other hand, the same AIHW report found that around 66% of women who were not pregnant met the WHO and Australian Department of Health guidelines (Australian Institute of Health and Welfare, 2019). A study based in Rockhampton Australia reported that 92% of study participants were not meeting the guidelines for exercise in pregnancy, but the study was limited by the small sample size (n = 142) and its regional location (Hayman et al., 2016), emphasising the need for further research on the Australian population. Collectively, this raises the question of why pregnant women are not meeting these guidelines. In the past, there have been concerns regarding the possible consequences of exercise in pregnancy on the foetus (Artal et al., 2003), however the current guidelines reflect that the many benefits outweigh any potential risks. Exercise has been shown to not be associated with a greater risk of miscarriage, perinatal mortality or birthing a child small for gestational age (Davenport et al., 2019a). A recently published Australian review on physical activity in pregnancy and the postpartum period reported no evidence of adverse outcomes of any form of exercise, including resistance training, during pregnancy (Brown et al., 2022). They further reported that physical activities during pregnancy should depend on individual preferences and pre-pregnancy routine activities (Brown et al., 2022). Furthermore, the review revealed a potential association between sedentary behaviour during pregnancy and poorer maternal and foetal circulatory health outcomes (e.g. venous pooling) (Brown et al., 2022). It is not clear if pregnant women are aware of the benefits of exercise and the guidelines regarding exercise in pregnancy, or whether they receive advice to exercise during their pregnancy. Insufficient advice from healthcare practitioners has been reported to contribute to the low rates of engagement in satisfactory exercise during pregnancy in USA, among Hispanic, African American, and White participants (Evenson et al., 2009), with the majority (60%) of American obstetricians indicating they were unaware of the recommended guidelines for exercising during pregnancy (McGee et al., 2018). A survey conducted in regional Australia (through 11 medical practices within the Rockhampton region of Central Queensland), reported that only 1.5% of respondents could recall a health practitioner providing them with exercise guidelines during pregnancy (Hayman et al., 2016). In 2015, Watson et al., reported in a South African convenience sample that despite the majority of medical practitioners believing that exercise in pregnancy was beneficial, the majority were not aware of the recommended guidelines and their advice did not always align with the current guidelines (Watson et al., 2015).

The aim of the current survey was to develop an understanding of Australian women’s perspectives and experiences around exercise during pregnancy, including women that experience depression or other mental illness. Specifically, this study aimed to obtain Australian women’s perspectives and experiences regarding:

1. Whether Australian women are aware of the national and international guidelines regarding exercise in pregnancy
2. Whether women, including those with depression or mental illness, were meeting these guidelines in pregnancy and what types of exercise women were performing
3. What advice women received about exercise during pregnancy, and
4. Women’s perception about the safety of exercise in pregnancy and the potential benefits of exercise in pregnancy for the prevention and management of depressive symptoms.

## Methodology

This survey study was approved by the University of Wollongong Human Research Ethics Committee (HREC) in compliance with the *National Statement on Ethical Conduct in Human Research* (2021/335).

### Study design

An anonymous online survey, administered via Qualtrics Australia, was used to collect data for this study. The survey aimed to develop an understanding of Australian women’s perspectives and experiences around exercise during pregnancy, including women that experience depression. It assessed women’s perspectives about: 1) safety of exercise during pregnancy, 2) preferable exercise type during pregnancy, 3) whether exercise in pregnancy is beneficial for depression/mood, 4) sources of medical advice about exercise in pregnancy and 5) whether women are meeting the WHO and the Australian Department of Health guidelines regarding exercise in pregnancy. The study population was defined as any person over the age of 18 who had previously been or were currently pregnant and was currently living in Australia. A subset of questions from the survey were directed at those participants who have never been pregnant. These questions and responses were excluded from the current project as they do not answer the aims of this study. Demographic details were collected followed by questions on the respondent’s knowledge and beliefs around the benefits and risks of exercise in pregnancy as well as their physical activity habits during pregnancy. The survey consisted of the following components:

1. Demographics
2. Pregnancy status (recent pregnancy, method of delivery, number of pregnancies, etc.)
3. Health status (including trusted medical information source, advice they received)
4. Attitudes/beliefs (regarding conducting exercise during pregnancy)
5. Physical activity habits during pregnancy

The final survey for those who have carried a pregnancy consisted of 45 questions. Response formats were varied to serve each question’s aim and type, to include fixed category format, free text responses and Likert scales. Skip logic resulted in having some questions not being displayed to all participants. Survey questions can be found in the supplementary materials.

### Validity of Questionnaire

The face validity of the survey was assessed by inviting 10 women of varying ages (≥18), including pregnant and non-pregnant women, to read aloud the questionnaire with a member of the research team. Participants then indicated their understanding and provided feedback to a member of the research team. Comments and suggestions were incorporated into the survey revision, to ensure that every question was clear and the format for answering was suitable.

### Pilot survey

The survey was piloted on the researchers’ personal social media networks for three days to improve reliability and validity for a wider audience. 47 participants responded to the pilot survey; analysis of this data resulted in several changes being made to the survey tool (e.g. inclusion of additional response options for some questions, correcting the survey logic).

### Recruitment

The survey was launched in November 2021 and remained open for four weeks. Invitation to participate in the survey was shared on various Facebook groups that were identified by the researchers to be of relevance to the target audience. Facebook groups were identified using key search terms such as “mums”, “bubs”, “baby”, “mothers”, “pregnant” and were restricted to Australian pages. The survey was also advertised on Twitter and Instagram.

### Data analysis

Data was analysed using IBM SPSS Statistics for Windows, Version 28.0. (IBM Corp., Armonk, NY, USA). Frequencies and descriptive statistics of respondent characteristics were calculated and presented in tables. Pearson’s Chi-Square analyses were performed to assess the association between variables according to prior mental health diagnosis, and pairwise z-tests adjusted to Bonferroni correction to assess the differences between columns’ proportions. Significance was reported when *p*<0.05.

## Results

### Demography

Demographic details are provided in Table 1. There was a total of 695 eligible responses. The majority (51%) of the participants were 26-35 years of age. Thirty-seven percent of respondents had been diagnosed with a mental illness during their life-time. Those that had been diagnosed with a mental illness during their lifetime had significantly more respondents in the 18-25 age category and fewer in the 46 and over category (*p*<0.001), fewer respondents with university level education (*p*<0.001) and lower household income (*p*<0.001). There was no difference in recency of pregnancy between those that had and had not been diagnosed with a mental illness during their lifetime, with 61% of all respondents being pregnant within the previous two years. One hundred and fifty-seven respondents (24%) reported experiencing mental illness during pregnancy; and 12% reported experiencing depression during pregnancy (Supp. Table S1).

**Table 1:** Demographic details of the responders.

| Variables n (%) |  |  | Diagnosed with a mental illness during their lifetime |  |  |  |
| --- | --- | --- | --- | --- | --- | --- |
|  |  | Total responses | Yes | No | Chi-square value | p-value |
| Age |  | 687 | 253 (36.8) | 434 (63.2) |  |  |
|  | 18-25 | 54 (7.9) | 31(12.3)* | 23(5.3)* | 23.773 | <0.001 |
|  | 26-35 | 349 (50.8) | 140(55.3) | 209 (48.2) |  |  |
|  | 36-45 | 205 (29.8) | 67 (26.5) | 138(31.8) |  |  |
|  | 46 and over | 79 (11.5) | 15(5.9)* | 64(14.7)* |  |  |
| State of residence |  | 683 | 252 (36.9) | 431 (63.1) |  |  |
|  | NSW | 458 (671) | 175 (69.4) | 283 (65.7) | 13.291 | 0.065 |
|  | ACT | 40 (5.9) | 17 (6.7) | 23 (5.3) |  |  |
|  | NT | 3 (0.4) | 0 (0) | 3 (0.7) |  |  |
|  | QLD | 69 (10.1) | 14 (5.6) | 55 (12.8) |  |  |
|  | SA | 15 (2.2) | 4 (1.6) | 11 (2.6) |  |  |
|  | TAS | 8 (1.2) | 4 (1.6) | 4 (0.9) |  |  |
|  | VIC | 61 (8.9) | 25 (9.9) | 36 (8.4) |  |  |
|  | WA | 29 (4.2) | 13 (5.2) | 16 (3.7) |  |  |
| Length of time since most recent pregnancy |  | 670 | 243 (36.3) | 427 (63.7) |  |  |
|  | Currently pregnant | 173 (25.8) | 59 (23.3) | 114 (26.7) | 3.103 | 0.541 |
|  | < 6 months | 102 (15.2) | 44 (18.1) | 58 (13.6) |  |  |
|  | 6-12 months | 65 (9.7) | 23 (9.5) | 42 (9.8) |  |  |
|  | 12-24 months | 66 (9.9) | 26 (10.7) | 40 (9.4) |  |  |
|  | > 2 years | 264 (39.4) | 91 (37.4) | 173 (40.5) |  |  |
| Experienced mental illness during pregnancy |  | 654 | 241 (36.9) | 413 (63.1) |  |  |
|  | No | 497(76) | 117(48.5)* | 380(92)* | 157.582 | <0.001 |
|  | Yes | 157(24) | 124(51.5)* | 33(8)* |  |  |
| Ethnicity |  | 683 | 253 (37) | 430 (63) |  |  |
|  | Non-Caucasian | 109 (16) | 35(13.8) | 74(17.2) | 1.353 | 0.245 |
|  | Caucasian | 574 (84) | 218(86.2) | 356(82.8) |  |  |
| Marital status |  | 688 | 254 (36.9) | 434 (63.1) |  |  |
|  | Married | 464 (67.4) | 141(55.5)* | 323(74.4)* | 32.196 | <0.001 |
|  | Cohabiting / Defacto | 154 (22.4) | 74(29.1)* | 80(18.4)* |  |  |
|  | Partner but live separately | 18 (2.6) | 11(4.3)* | 7(1.6)* |  |  |
|  | Divorced | 21 (3.1) | 8(3.1)* | 13(3)* |  |  |
|  | Single | 26 (3.8) | 16(6.3)* | 10(2.3)* |  |  |
|  | Other | 1 (0.1) | 1(0.4) | 0(0.0) |  |  |
|  | Prefer not to say | 4 (0.6) | 3(1.2) | 1(0.2) |  |  |
| Education level |  | 687 | 254 (37) | 433 (63) |  |  |
|  | Less than high school | 22 (3.2) | 9(3.5) | 13(3) | 24.468 | <0.001 |
|  | HSC or year 12 equivalent | 56 (8.2) | 31(12.2)* | 25(5.8)* |  |  |
|  | Certificates I-V (TAFE) | 97 (14.1) | 49 (19.3)* | 48 (11.1)* |  |  |
|  | Diploma | 81 (11.8) | 31(12.2) | 50(11.5) |  |  |
|  | (Diploma and above) # | 256 (37.3) | 120(47.2)* | 136(31.4)* |  |  |
|  | Undergraduate degree | 226 (32.9) | 63(24.8)* | 163(37.6)* |  |  |
|  | Postgraduate degree | 205 (29.8) | 71(28) | 134(30.9) |  |  |

| Employment Status<sup>(a)</sup> |  |  |  |  |  |  |
| --- | --- | --- | --- | --- | --- | --- |
|  | Student | 45 (6.5) | 23 (9.1) | 22 (5.1) | 35.155 | <0.001 |
|  | Unemployed | 66 (9.5) | 33 (13.1)* | 33 (7.6)* |  |  |
|  | Maternity leave | 140 (20.1) | 58 (23) | 82 (18.9) |  |  |
|  | Casual job | 51 (7.3) | 22 (8.7) | 29 (6.7) |  |  |
|  | Full-time job | 253 (36.4) | 65 (25.8)* | 188 (43.3)* |  |  |
|  | Part-time job | 168 (24.2) | 67 (26.6) | 101 (23.3) |  |  |
|  | Retired | 12 (1.7) | 3 (1.2) | 9 (2.1) |  |  |
| Household Income |  | 685 | 252 (36.8) | 433 (63.2) |  |  |
| | < \$18,200 | 6 (0.9) | 3(1.2) | 3(0.7) | 31.374 | <0.001 |
| | \$18,201 - \$37,000 | 26 (3.8) | 16(6.3)* | 10(2.3)* | | |
| | \$37,001 - \$80,000 | 76 (11.1) | 44(17.5)* | 32(7.4)* | | |
| | > \$80,001 | 493 (72) | 162(64.3)* | 331(76.4)* | | |
|  | Prefer not to say | 50 (7.3) | 11(4.4)* | 39(9)* |  |  |
|  | Not applicable | 34 (5) | 16(6.3) | 18(4.2) |  |  |

ACT: Australian Capital Territory, NSW: New South Wales, NT: North Territory, QLD: Queensland, SA: South Australia, TAS: Tasmania, VIC: Victoria, WA: Western Australia, <sup>(a)</sup>: Multiple responses Chi-square analysis, <sup>#</sup>: collapsed category comprised of diploma, undergraduate degree and postgraduate degree, and was not run in a separate analysis, \* signifies the pairwise comparisons. Responses are presented as n (%) No question was compulsory to answer, therefore the total number of responders for each question may vary.

### Awareness of WHO and Australian guidelines

Descriptive analysis showed that 425 (71%) of the responders were not aware of the WHO and Australian Government Department of Health guidelines regarding exercise during pregnancy, and this findings was not influenced by mental health diagnosis (Table 2). Moreover, education level did not influence the number of the participants who were aware of the WHO and Australian Department of Health guidelines.

**Table 2:**
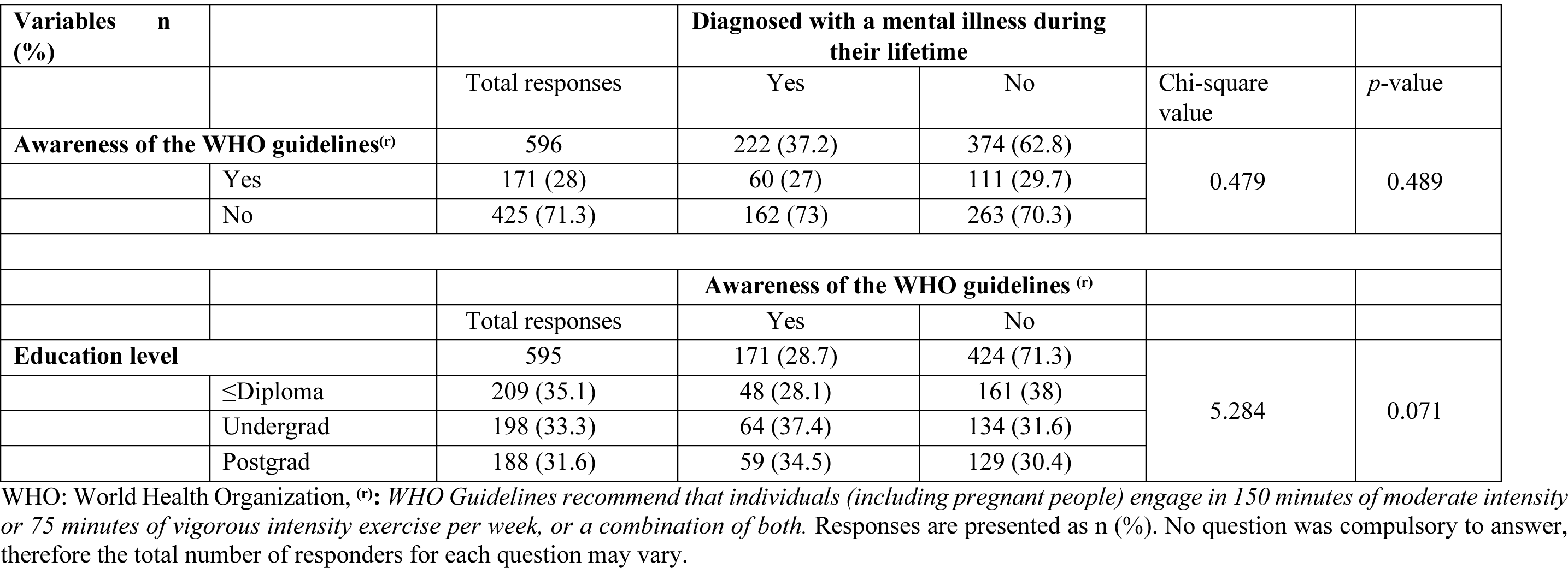
Awareness of WHO and Australian Health guidelines.

### Perspectives on safety and benefits of exercise during pregnancy

Descriptive analysis showed the majority of responders (94%) agree that exercise during pregnancy is safe for both mother and baby (Table 3), with 88-96% agreeing it is safe during each of the three trimesters (Supp. Table S1). There was a significant association between a mental illness diagnosis and the belief that exercise is safe for the mother and baby (X2 = 12.442, *p* = 0.014). Pairwise comparisons revealed that those with a mental illness diagnosis were less likely to strongly agree, and more likely to be neutral with this statement. Eighty-eight percent of responders agree that regular exercise would help to prevent a decline in a pregnant person’s mood and 92% agree that regular exercise would help to improve a pregnant person’s mood (Table 3). The majority of responders (73%) believed that pregnant women should preform 150 mins or less of moderate intensity exercise/week for general health and most responders believed that pregnant women should perform 60 mins or less of vigorous intensity exercise/week for general health (Table 3). These perspectives were not influenced by a mental health diagnosis (Table 3, *p* > 0.05).

**Table 3:**
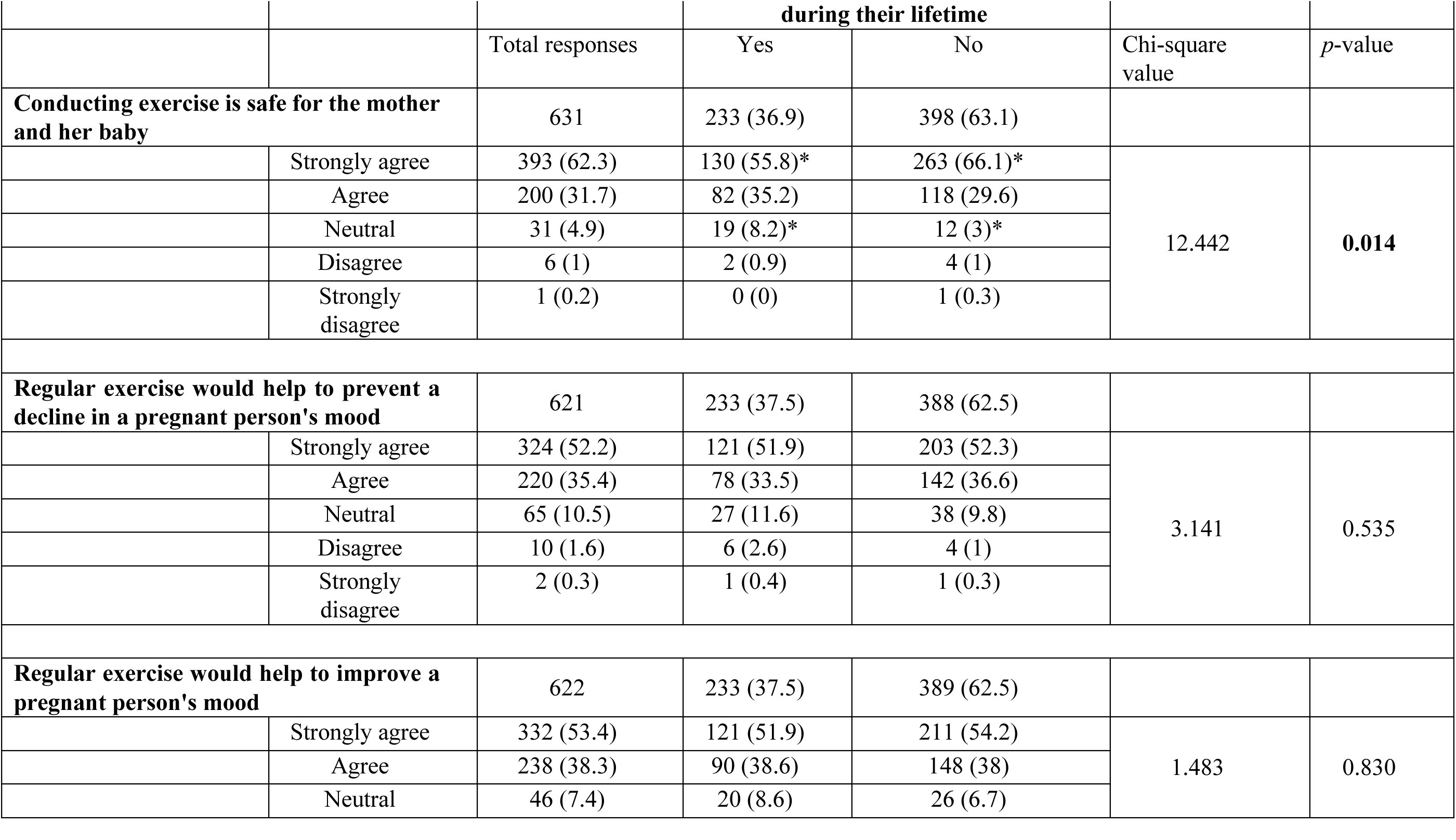
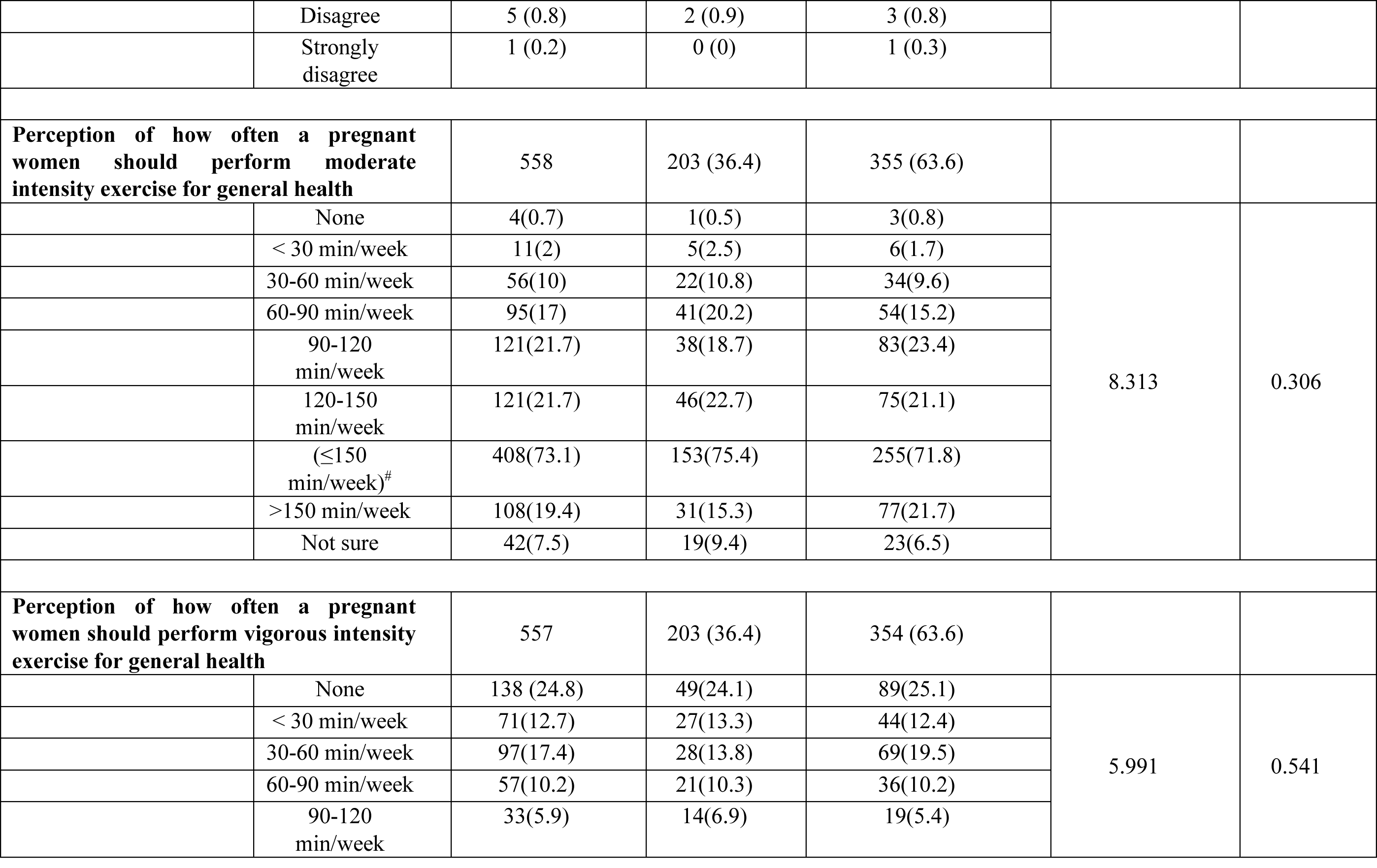
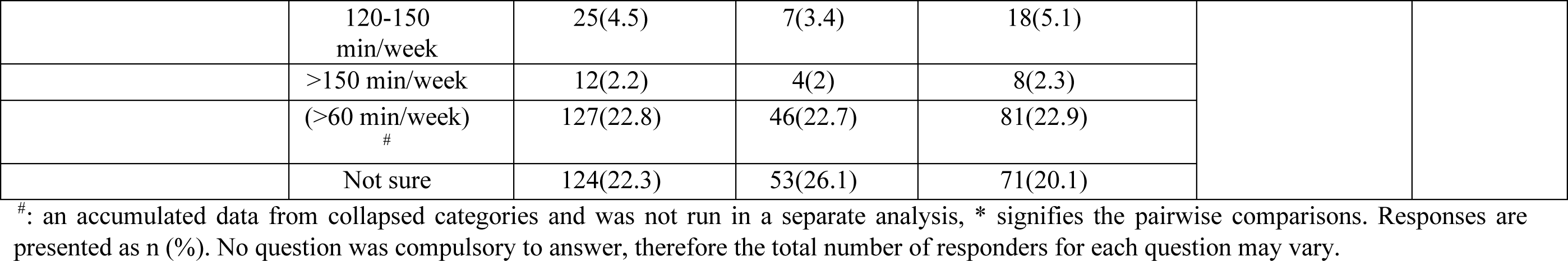
Participants’ beliefs and perspectives toward exercise during pregnancy.

### Type and source of medical advice/information received about exercise during pregnancy

Data related to advice participants received about exercising during pregnancy is provided in Table 4. Descriptive analysis showed that 71% of respondents considered GPs to be the most widely considered as a trusted source of medical advice including during pregnancy. Fifty-seven percent of responders recall being advised to exercise during their most recent pregnancy, mostly by their GP (54%), midwife (46%) and obstetrician (38%). Walking was the most advised exercise type (80%) followed by swimming (46%). Overall, 69% of responders report receiving no or little advice from their healthcare provider about the safety and benefits of exercise during their most recent pregnancy, but 45% of responders felt satisfied about the advice they did receive.

**Table 4:** Type and source of provided medical advice/information about exercise during pregnancy:

| Variables n (%) |  |  | Diagnosed with a mental illness during their lifetime |  |  |  |
| --- | --- | --- | --- | --- | --- | --- |
|  |  | Total responses | Yes | No | Chi-square value | p-value |
| Trusted medical resource<sup>(a)</sup> |  |  |  |  |  |  |
|  | GP | 491 (70.6) | 189 (75.3) | 302 (70.1) | 70.446 | <0.001 |
|  | Midwife | 364 (52.4) | 137 (54.6) | 227 (52.7) |  |  |
|  | Obstetrician/Gynaecologist | 326 (46.9) | 115 (45.8) | 211 (49) |  |  |
|  | Friends/ family | 123 (17.7) | 45 (17.9) | 78 (18.1) |  |  |
|  | Psychiatrist/psychologist | 63 (9.1) | 51 (20.3)* | 12 (2.8)* |  |  |
|  | Online media | 48 (6.9) | 14 (5.6) | 34 (7.9) |  |  |
|  | Cultural medicine | 19 (2.7) | 8 (3.2) | 11 (2.6) |  |  |
|  | Physiotherapist/ EP/Chiro | 14 (2) | 4 (1.6) | 10 (2.3) |  |  |
|  | Scientific/medical sources/govt. websites | 11 (1.6) | 6 (2.4) | 5 (1.2) |  |  |
|  | Other | 19 (2.7) | 2 (0.8)* | 17 (3.9)* |  |  |
| Were responders advised to exercise during their most recent pregnancy |  | 681 | 250 (36.7) | 431 (63.3) |  |  |
|  | Yes | 386 (56.7) | 141 (56.4) | 245 (56.8) | 0.039 | 0.981 |
|  | No | 185(27.2) | 69(27.6) | 116(26.9) |  |  |
|  | Unsure | 110(16.2) | 40(16) | 70(16.2) |  |  |
| Source of that advice<sup>(a)</sup> |  |  |  |  |  |  |
|  | GP | 201 (53.5) | 83 (61) | 118 (49.2) | 8.628 | 0.196 |
|  | Midwife | 174 (46.3) | 67 (49.3) | 107 (44.6) |  |  |
|  | Obstetrician | 141 (37.5) | 49 (36) | 92 (38.3) |  |  |
|  | Family/Friend | 59 (15.7) | 25 (18.4) | 34 (14.2) |  |  |
|  | Personal trainer/Gym instructor | 47 (12.5) | 15 (11) | 32 (13.3) |  |  |
|  | Other | 48 (12.8) | 14 (10.3) | 34 (14.2) |  |  |
| Type of exercise that was advised during pregnancy<sup>(a)</sup> |  |  |  |  |  |  |
|  | Walking | 301 (80.1) | 112 (82.4) | 189 (78.8) | 10.64 | 0.301 |
|  | Swimming | 174 (46.3) | 66 (48.5) | 108 (45) |  |  |
|  | Yoga | 103 (27.4) | 44 (32.4) | 59 (24.6) |  |  |
|  | Weight Lifting/Resistance | 84 (22.3) | 27 (19.9) | 57 (23.8) |  |  |
|  | No specific | 59 (15.7) | 20 (14.7) | 39 (16.3) |  |  |
|  | Running | 40 (10.6) | 10 (7.4) | 30 (12.5) |  |  |
|  | Cycling | 26 (6.9) | 7 (5.1) | 19 (7.9) |  |  |
|  | Other | 62 (16.5) | 17 (12.5) | 45 (18.8) |  |  |
|  | Cannot remember | 5 (1.3) | 2 (1.5) | 3 (1.3) |  |  |
| Amount of advice received by healthcare provider(s) about exercise safety and benefits during pregnancy |  | 665 | 243 (36.5) | 422 (63.5) |  |  |
|  | None at all | 170(25.6) | 56(23) | 114(27) | 3.513 | 0.621 |
|  | A little | 288(43.3) | 113(46.5) | 175(41.5) |  |  |
|  | A moderate amount | 134(20.2) | 44(18.1) | 90(21.3) |  |  |
|  | A lot | 23(3.5) | 10(4.1) | 13(3.1) |  |  |
|  | A great deal | 19(2.9) | 7(2.9) | 12(2.8) |  |  |
|  | Unsure | 31(4.7) | 13(5.3) | 18(4.3) |  |  |

| **Level of satisfaction with advice received from their healthcare provider regarding the safety & benefits of exercise during pregnancy** |  | 663 | 243 (36.7) | 420 (63.3) |  |  |
| --- | --- | --- | --- | --- | --- | --- |
|  | Very satisfied | 97(14.6) | 34(14) | 63(15) | 7.785 | 0.100 |
|  | Satisfied | 202(30.5) | 60(24.7) | 142(33.8) |  |  |
|  | Neutral | 281(42.4) | 118(48.6) | 163(38.8) |  |  |
|  | Dissatisfied | 62(9.4) | 23(9.5) | 39(9.3) |  |  |
|  | Very dissatisfied | 21(3.2) | 8(3.3) | 13(3.1) |  |  |

<sup>(a)</sup>: Multiple responses Chi-square analysis. Responses are presented as n (%). No question was compulsory to answer, therefore the total number of responders for each question may vary.

### Exercise during pregnancy

Participant responses regarding the amount of and type of exercise conducted during pregnancy are shown in Table 5. Descriptive analysis showed that 412 (68%) out of 610 responders conducted exercise during all or part of their most recent pregnancy. There was a significant association between a reported diagnosis of a mental illness during their lifetime and participation in structured exercising during pregnancy (X^2^=6.862, *p*=0.009), with fewer individuals with a mental illness exercising during pregnancy compared to those without (Table 5). Similarly, there was a significant association between a reported diagnosis of a mental illness during their lifetime and participation in structured exercising prior to pregnancy (X^2^=10.225, *p*=0.001), with fewer individuals with a mental illness participating in exercise prior to pregnancy. Of those respondents that reported participating in structured exercise during all/part of their most recent pregnancy, 51% reported exercising 3-4 times per week and 23% less than 3 times/week; 79% reported conducting less than or equal to 150mins of moderate intensity exercise/week and 78% reported conducting 60mins and less of vigorous intensity exercise/week. There was no significant association between mental illness diagnosis during their lifetime and the frequency nor total time spent in conducting exercise during pregnancy (p > 0.05). The most common types of exercise conducted during pregnancy were walking (87%), swimming (38%) and yoga (36%) (Table 5). However there was a significant association between mental illness diagnosis and type of exercise conducted during pregnancy with fewer respondents with a mental illness performing higher intensity exercises such as swimming, running, cyclising HIIT and more yoga (Table 5).

**Table 5:** Exercise experience during the most recent pregnancy.

| Variables n (%) |  |  | Diagnosed with a mental illness during their lifetime |  |  |  |
| --- | --- | --- | --- | --- | --- | --- |
|  |  | Total responses | Yes | No | Chi-square value | p-value |
| Participating structured exercise during all/part of pregnancy |  | 610 | 229 (37.5) | 381 (62.5) |  |  |
|  | Yes | 412 (67.5) | 140(61.1)* | 272(71.4)* | 6.862 | **0.009** |
|  | No | 198 (32.5) | 89(38.9)* | 109(28.6)* |  |  |
| Practiced exercise prior to falling pregnant, or during previous pregnancies |  | 567 | 216 (38.1) | 351 (61.9) |  |  |
|  | Yes | 472 (83.2) | 166(76.9)* | 306(87.2)* | 10.225 | **0.001** |
|  | No | 95 (16.8) | 50(23.1)* | 45(12.8)* |  |  |
| Pregnancy trimester and participation in exercise |  |  |  |  |  |  |
| 1st trimester |  | 398 | 132 (33.2) | 266 (66.8) |  |  |
|  | Yes | 350 (87.9) | 108 (81.8)* | 242 (91*) | 7.838 | **0.020** |
|  | No | 42 (10.6) | 20 (15.2)* | 22 (8.3)* |  |  |
|  | Unsure | 6 (1.5) | 4 (3) | 2 (0.8) |  |  |
| 2nd trimester |  | 393 | 131 (33.3) | 262 (66.7) |  |  |
|  | Yes | 367(93.4) | 118 (90.1) | 249 (95) | 4.293 | 0.117 |
|  | No | 18(4.6) | 8(6.1) | 10(3.8) |  |  |
|  | Unsure | 8(2) | 5(3.8) | 3(1.1) |  |  |
| 3rd trimester |  | 381 | 128 (33.6) | 253 (66.4) |  |  |
|  | Yes | 300(78.7) | 102(79.7) | 198(78.3) | 0.798 | 0.671 |
|  | No | 55(14.4) | 16(12.5) | 39(15.4) |  |  |
|  | Unsure | 26(6.8) | 10(7.8) | 16(6.3) |  |  |
| Frequency of exercise during most recent pregnancy |  | 401 | 136 (33.9) | 265 (66.1) |  |  |
|  | <3 times a week | 94(23.4) | 36(26.5) | 58(21.9) | 1.281 | 0.530 |
|  | 3-4 times a week | 203(50.6) | 68(50) | 135(50.9) |  |  |
|  | ≥5 times a week | 104(25.9) | 32(23.5) | 72(27.2) |  |  |
| Total time spent conducting moderate physical activity/week |  | 401 | 136 (33.9) | 265 (66.1) |  |  |
|  | None | 11(2.7) | 6(4.4) | 5(1.9) | 9.566 | 0.215 |
|  | <30 min/week | 19(4.7) | 10(7.4) | 9(3.4) |  |  |
|  | 30-60 min/week | 49(12.2) | 16(11.8) | 33(12.5) |  |  |
|  | 60-90 min/week | 83(20.7) | 33(24.3) | 50(18.9) |  |  |
|  | 90-120 min/week | 89(22.2) | 29(21.3) | 60(22.6) |  |  |
|  | 120-150 min/week | 64(16) | 20(14.7) | 44(16.6) |  |  |
|  | (≤150min) # | 315(78.6) | 114(83.8) | 201(75.8) |  |  |
|  | >150 min/week | 77(19.2) | 19(14) | 58(21.9) |  |  |
|  | Unsure | 9(2.2) | 3(2.2) | 6(2.3) |  |  |
| Total time spent conducting vigorous physical activity/week |  | 402 | 136 (33.8) | 266 (66.2) |  |  |
|  | None | 150(37.3) | 56 (37.3) | 94 (62.7) | 3.745 | 0.809 |
|  | <30 min/week | 64(15.9) | 21(15.4) | 43(16.2) |  |  |
|  | 30-60 min/week | 87(21.6) | 27(19.9) | 60(22.6) |  |  |
|  | 60-90 min/week | 41(10.2) | 16(11.8) | 25(9.4) |  |  |
|  | 90-120 min/week | 24 (6) | 6(4.4) | 18(6.8) |  |  |
|  | 120-150 min/week | 13 (3.2) | 4 (2.9) | 9 (3.4) |  |  |
|  | >150 min/week | 12 (3) | 4 (2.9) | 8 (3) |  |  |
|  | (≥60 min) # | 90 (22.4) | 30 (22.1) | 60 (22.6) |  |  |
|  | Unsure | 11 (2.7) | 2 (1.5) | 9 (3.4) |  |  |
| Type of exercise conducted during pregnancy<sup>(a)</sup> |  |  |  |  |  |  |
|  | Aerobic | 381 (94.5) | 127 (93.4) | 254 (95.1) | 6.095 | 0.192 |
|  | resistance | 171 (42.4) | 53 (39) | 118 (44.2) |  |  |
|  | Both | 174 (43.2) | 56 (41.2) | 118 (44.2) |  |  |
|  | Yoga/Pilates | 170 (42.2) | 67 (49.3) | 103 (38.6) |  |  |
|  | Walking | 350 (86.8) | 121 (89) | 229 (86.7) | 28.935 | **0.001** |
|  | Swimming | 153 (38) | 38 (27.9)* | 115 (43.6)* |  |  |
|  | Yoga | 143 (35.5) | 59 (43.4)* | 84 (31.8)* |  |  |
|  | Running/Juggling | 117 (29) | 33 (24.3)* | 84 (31.8)* |  |  |
|  | HITT/Cross fit | 75 (18.6) | 22 (16.2)* | 53 (20.1)* |  |  |
|  | Cycling | 65 (16.1) | 14 (10.3)* | 51 (19.3)* |  |  |
|  | Pilates | 37 (9.2) | 11 (8.1) | 26 (9.8) |  |  |
|  | Team support | 17 (4.2) | 5 (3.7) | 12 (4.5) |  |  |
|  | Aerobics/Dance | 31 (7.7) | 16 (11.8) | 15 (5.7) |  |  |
|  | Martial arts combat | 4 (1) | 1 (0.7) | 3 (1.1) |  |  |
| Reason for stopping exercise during pregnancy<sup>(a)</sup> |  |  |  |  |  |  |
|  | Feeling fatigue | 119 (61.7) | 54 (62.8) | 65 (60.7) | 10.231 | 0.176 |
|  | Morning sickness | 105 (54.4) | 54 (62.8) | 51 (47.7) |  |  |
|  | Not enough time | 76 (39.4) | 36 (41.9) | 40 (37.4) |  |  |
|  | Concerns about the baby safety | 26 (13.5) | 9 (10.5) | 17 (15.9) |  |  |
|  | Concerns about self-safety | 17 (8.8) | 5 (5.8) | 12 (11.2) |  |  |
|  | Medical advice | 14 (7.3) | 9 (10.5) | 5 (4.7) |  |  |
|  | Other | 57 (29.5) | 26 (30.2) | 31 (29) |  |  |

<sup>(a)</sup>: Multiple responses Chi-square analysis, #: an accumulated data from collapsed categories and was not run in a separate analysis, \*: signifies the pairwise comparisons. Responses are presented as n (%). No question was compulsory to answer, therefore the total number of responders for each question may vary

### Participants who experienced mental illness during pregnancy (including antenatal depression)

Twenty four percent of survey responders reported experiencing a mental illness during their most recent pregnancy (Table 1), including 12% depression, 21% anxiety, and 1% other, with some respondents experiencing more than one mental illness (Supp. Table S1). Fifty-three percent of the responders who experienced mental illness during their most recent pregnancy were advised to conduct exercise, similar to those that did not experience a mental illness (Supp. Table S3). Interestingly, out of the 157 responders who experienced mental illness during their pregnancy, 72% report receiving little-to-no advice about the safety and benefits of conducting exercise during pregnancy (Supp. Table S3). Seventy one percent of the responders who had not experienced mental illness during their most recent pregnancy reported that they conducted exercise during their most recent pregnancy (Supp. Table S3). This was significantly different in those that experienced mental illness during pregnancy, with 57% conducting exercise during their most recent pregnancy (X^2^= 8.306, *p*=0.004) (Supp. Table S3), specifically this significant difference was during the first and third pregnancy trimester (Supp. Table S3). Only 164 (29%) of the 570 responders were aware of the WHO and the Australian Department of Health guidelines regarding exercise during pregnancy; and there was no significant association between this awareness and an experience of mental illness during pregnancy (Supp. Table S3).

There was a significant association between a reported experience of a mental illness during pregnancy and conducted exercise type during pregnancy (X^2^=21.326, *p*=0.019) (Supp. Table S3). Although walking was the most common conducted exercise during pregnancy in those with and without a mental illness during pregnancy, fewer respondents with a mental illness during pregnancy reported swimming, running/jugging and cycling during their pregnancies, while more women with a mental illness during pregnancy reported practicing yoga during their pregnancies (Supp. Table S3). There was also a significant association between experiencing mental health illness during pregnancy and the belief that exercise is safe for the mother and baby (X2 = 16.191, *p* = 0.003), with fewer responders experiencing mental health illness during pregnancy who strongly agree that exercise is safe for the mother and baby (Supp. Table S3).

## Discussion

Evidence suggests that exercise during pregnancy can have a positive influence on maternal physical and mental health, as well as newborn health outcomes. Accordingly, the WHO and National guidelines recommend women exercise during pregnancy. Our study showed that most women surveyed were not aware of the WHO/National guidelines regarding exercise in pregnancy. Despite this, the vast majority believed that regular exercise during pregnancy is safe for mother and baby and that it would help prevent a decline in and improve a pregnant person’s mood. Most of the responders also reported conducting exercise during all or part of their most recent pregnancy; however, there were significantly fewer individuals with a mental illness that reported exercising during pregnancy. Despite the majority participating in exercise during pregnancy, more than half of responders report conducting less exercise than that recommended by WHO. While just over half of the responders recall being advised to exercise during their most recent pregnancy, the majority report receiving no or little advice from their healthcare provider about the safety and benefits of exercise during their most recent pregnancy.

### Knowledge and beliefs about exercise in pregnancy

The current survey showed that the majority of responders were not aware of the WHO and Australian Health guidelines, in regard to conducting exercise during pregnancy. To our knowledge the current study is the first to assess the awareness of Australian women about the WHO and Australian Health guidelines, in regard to exercise during pregnancy. Notwithstanding, despite the lack of knowledge of the existing international/national guidelines towards exercise during pregnancy, the majority believe that exercise during pregnancy is safe and beneficial for a person’s mood, specifically. In line with these findings, a previous study, conducted in Victoria Australia reported that the majority believed that regular low intensity exercise was safe during pregnancy; while the majority also believed moderate intensity exercise use was safe, only a small percentage believed high intensity exercise was safe during pregnancy (Duncombe et al., 2009). Similarly, our findings showed that many responders believe women should be performing no or limited vigorous activity per week.

The current finding, regarding the belief of safe and beneficial of exercise during pregnancy, was consistent with several international surveys that were spread among their general population. For example, in a USA study, it was reported that among the 500 participants there were 76% who believe that exercise during pregnancy is safe and 86% believe it is beneficial (Babbar and Chauhan, 2015). Similarly, in a study from Saudi Arabia, out of the 472 participants 67% believed that exercise during pregnancy is safe (Al-Youbi and Elsaid, 2020). However in Ethiopia, only 52% of the 403 participants believed that exercise is safe during pregnancy (Janakiraman et al., 2021). A theoretic explanation for this drop in the percentage from USA to Saudi to Ethiopia, might be as a result of the different cultures, as it was previously reported in both Saudi Arabian and Ethiopian reports that the majority of pregnant women believe that physical exercise would cause a miscarriage, early delivery and low birth weight, also it might be harmful to their health during pregnancy (Addis et al., 2022; Gari et al., 2022; Hjorth et al., 2012; Musaiger et al., 2012); however this belief has recently started to change (Addis et al., 2022; Gari et al., 2022).

An overwhelming majority of responders report receiving no or little advice from their healthcare provider about the safety or benefits of exercise during their most recent pregnancy, and this did not differ between those that have and have not been diagnosed with a mental illness. Our finding was consistent with a Chinese survey, that reported only 8% of pregnant women have received advice from a medical practitioner about the safety and benefits of physical activity during pregnancy (Zhang et al., 2014). Furthermore, only 33% of Ethiopian pregnant women were advised to conduct exercise during their pregnancy, according to a recent survey (Janakiraman et al., 2021). This does raise a question about whether health care providers have enough knowledge and awareness of the safety and benefits of exercise during pregnancy. To our knowledge, there are no Australian studies that have assessed medical practitioners’ knowledge towards the WHO guidelines regarding exercise during pregnancy, however there are several international assessments. In Michigan, USA, 60% of practicing healthcare providers and 86% of physicians and certified nurse midwives were not familiar with the guidelines (Bauer et al., 2010). Among health care providers in South Africa, 83% were unaware of the WHO guidelines (Watson et al., 2015), while in Pakistan 72% of the medical practitioners were unaware of the WHO guidelines towards exercise during pregnancy (Batool et al., 2021). Collectively, this suggests greater international effort is needed to educate health care providers so they can provide appropriate guidance to their patients.

Exercise conducted during pregnancy The majority of women surveyed in the current study report they did conduct exercise during their most recent pregnancy. Of those that exercised, many did not meet the WHO guidelines. This current finding was consistent with the Australian Institute of Health and Welfare report in 2019 that stated: “two-thirds of pregnant women were active for fewer than 150 minutes per week” (Australian Institute of Health and Welfare, 2019). Also, Zhang et al., have reported that only 11% of the pregnant women have managed to meet with the WHO guidelines (Zhang et al., 2014). This finding is also consistent with reports from other countries including Ethiopia (Janakiraman et al., 2021), Saudi Arabia (Al-Youbi and Elsaid, 2020), USA (specifically Virginia state) (Babbar and Chauhan, 2015) and China (Zhang et al., 2014), with reports in these studies of 11-23% of women meeting the WHO guidelines in pregnancy. We found that significantly fewer women diagnosed with a mental illness, including those with antenatal depression, report performing exercise in pregnancy. This is consistent with an Australian pregnancy cohort study (Watson et al., 2018) that reported an increase in depressive and anxiety symptoms was associated with a steeper decline in exercise frequency during the perinatal period, across pregnancy trimesters and postpartum period (up to 12 months after delivery) (Watson et al., 2018). Interestingly, Watson et al., reported that women with depression and taking antidepressant medication conducted significantly less exercise compared to the control group, during the first trimester (Watson et al., 2018). According to a growing body of literature, exercise has been shown to bring positive effects to mood states such as anxiety, stress, and depression (in non-pregnant populations) (Cooney et al., 2013; El-Kade and Al-Jiffri, 2016; López-Torres Hidalgo et al., 2019), with emerging evidence suggesting this also extends to the pregnant population (Jarbou and Newell, 2022).

Understanding the barriers to conducting exercise during pregnancy will assist in developing strategies to support women to meet the WHO guidelines. In this study, the main barriers to exercise in pregnancy reported by Australian women were fatigue, morning sickness, and lack of time; and there were no significant differences in the barriers reported between those with and without a diagnosed mental illness in their lifetime. Interestingly, barriers appear to have changed over time, as according to a Victorian survey in 2009, in Australia, the concerns of pregnancy safety was one of the major barriers to conduct exercise during pregnancy, followed by feeling tired (Duncombe et al., 2009). Internationally, some of the main barriers to conduct exercise during pregnancy were similar, for example, in Saudi-Arabia 84% of women reported feeling tired was the main barrier to conducting exercise during pregnancy, followed by a lack of knowledge of the importance of maternal exercise, and a lack of time and transportation difficulties (Al-Youbi and Elsaid, 2020). However, barriers to conduct exercise during pregnancy were different in both the Ethiopian and Chinese studies, where the main barrier was a concern for the foetus safety (78% and 67% of the participants, respectively), followed by a lack of knowledge of the WHO guidelines regarding exercise during pregnancy (Guelfi et al., 2015; Janakiraman et al., 2021; Zhang et al., 2014). This highlights some important varieties about the beliefs and attitudes towards exercise during pregnancy among different cultures. This was previously reported by Guelfi et al., that compared Chinese to Australian pregnant women’s beliefs about exercise during pregnancy (Guelfi et al., 2015). Their study reported that Australian pregnant women conducted significantly more vigorous and moderate-intensity physical activity compared to the Chinese women, while Chinese women reported more walking activity than the Australian women (Guelfi et al., 2015). However, there was no significant difference between groups when the Metabolic equivalent minutes (MET-minute) per week were calculated for all activities combined (Guelfi et al., 2015). Differences between cultures, beliefs and attitudes need to be fully explored in future studies to assist in the design of exercise interventions to maximise exercise adherence and lifelong activity behaviour.

The most common types of exercise that the responders report conducting during their most recent pregnancy were walking, swimming, then yoga, which was similar to what they report being generally advised to conduct during their most recent pregnancy. This was consistent with the Virginia, USA study (Babbar and Chauhan, 2015), as almost 62% conducted walking, 20% swimming and 15% were practicing yoga during pregnancy (Babbar and Chauhan, 2015). Walking was the most conducted exercise during pregnancy in China, Ethiopia and Saudi Arabia (98%, 91% and 66%, respectively (Al-Youbi and Elsaid, 2020; Janakiraman et al., 2021; Zhang et al., 2014). These exercises would be considered as low intensity exercises. It is possible that the focus on low intensity exercise is derived from a lack of knowledge about the types of exercise and safety of different exercises in pregnancy. As discussed by Watson et al, 2015, clinicians may see pregnancy as a high-risk state which leads to low intensity exercise such as walking, yoga and swimming being the most highly recommended (Watson et al., 2015).

### Study strengths and limitations

Despite the fact that similar studies have been conducted in other countries (Al-Youbi and Elsaid, 2020; Janakiraman et al., 2021; Zhang et al., 2014), this study is the first of its kind assessing Australian women’s beliefs and attitudes towards exercise during pregnancy, as well as their levels of satisfaction with the advice provided regarding exercise during pregnancy. Such research is essential to determine the next step in implementing effective protocols to raise awareness of WHO guidelines for exercise during pregnancy, and how essential, beneficial and safe is exercise during pregnancy. However, there were several limitations of this study which need to be considered.

1. Online surveys of this nature are limited to responders that have internet access and likely who were interested in the topic (Andrade, 2020) and due to the unrestricted selection of participants, voluntary online surveys have limited generalizability to the target population (Singh and Sagar, 2021).
2. This survey was only distributed in English, limiting responses to those that understand English. It is estimated that approximately 2% of the Australian population do not speak English (Australia Community Profile, 2021); it may be that these communities have different perceptions and experiences regarding exercise during pregnancy.
3. Australia is considered a multi-cultural society (Australian Human Rights Commission, 2022), however, the majority of responders were Caucasian (84%, Table 1). This limits the perceptions of different cultural or ethnic groups. As based on previously discussed evidence, experience, believes and barriers regarding exercise during pregnancy could be different between different cultures.
4. This survey required responders to recall exercise amount and advice received from several years prior, with 40% of responders required to recall from more than two years prior, subjecting responses to recall bias. Prospectively tracking exercise conducted during pregnancy would provide a more accurate indication.
5. The majority of the participants that responded were from NSW, and in the 26-45 age range (Table 1), biasing responses towards this demographic. While the available evidence shows that surveys conducted in different regions/states in Australia report similar findings to the present study (Hayman et al., 2016; Newham et al., 2016), it is expected that the under 25 and 45+ demographic may have differing views and experiences.

## Conclusion

This study has shown that Australian women believe that exercise during pregnancy is safe and has benefits for mental health. However, many women report receiving little advice about this from their healthcare provider. Furthermore, women report not being aware of or meeting the WHO/National guidelines regarding exercise in pregnancy. Women do report primarily conducting low intensity exercise during pregnancy (walking, swimming, yoga). Importantly, fewer respondents with a diagnosed mental illness report exercising during pregnancy. Considering the potential benefits of exercise for mental illness, employing strategies to increase this group of women’s engagement with exercise during pregnancy is important. The main barriers to exercise include fatigue and time. Further research to determine strategies to increase exercise in pregnant women, particularly those with a mental illness, are needed.

## Data Availability

All data produced in the present study are available upon reasonable request to the authors

## Supplementary Material

**Table S1:**
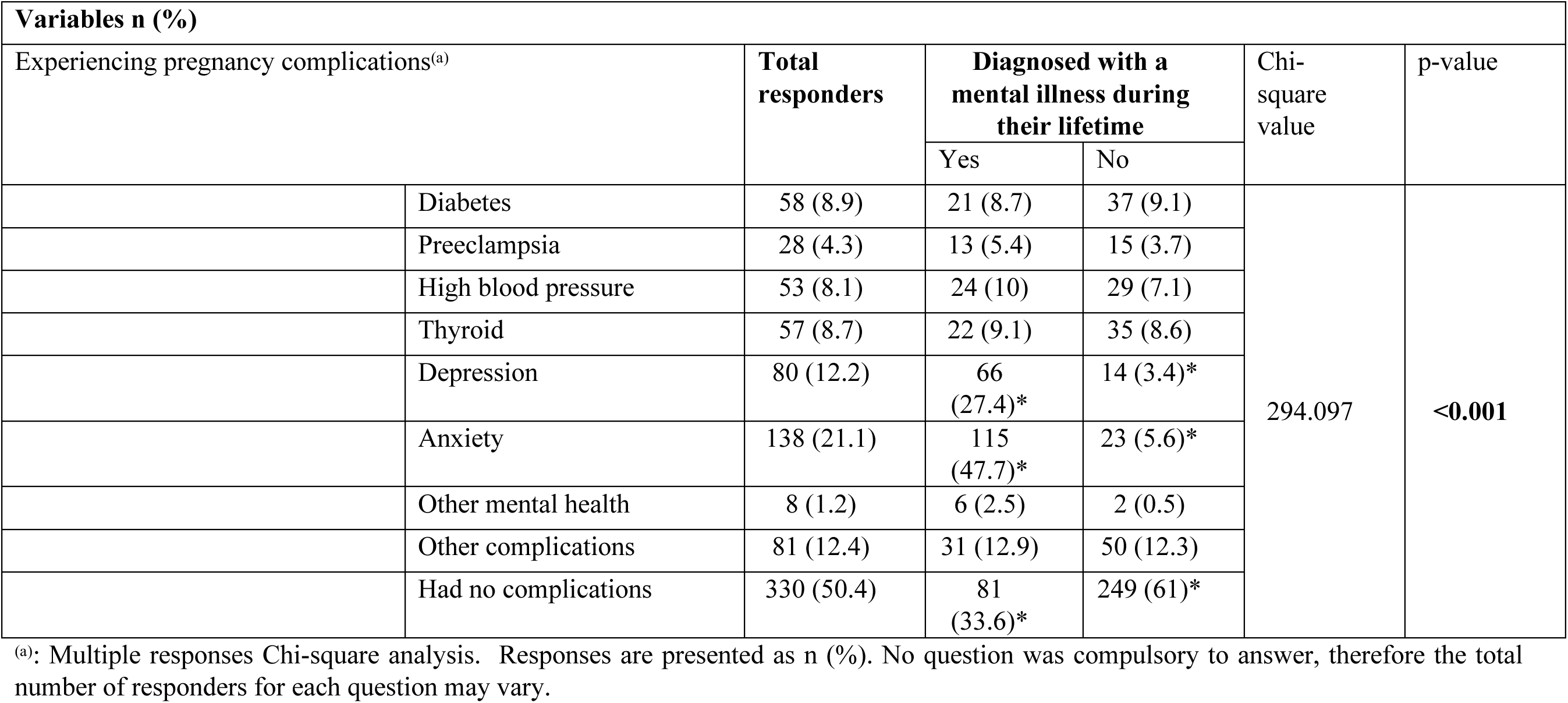
What medical conditions did you experienced during your most recent pregnancy?

**Table S2:**
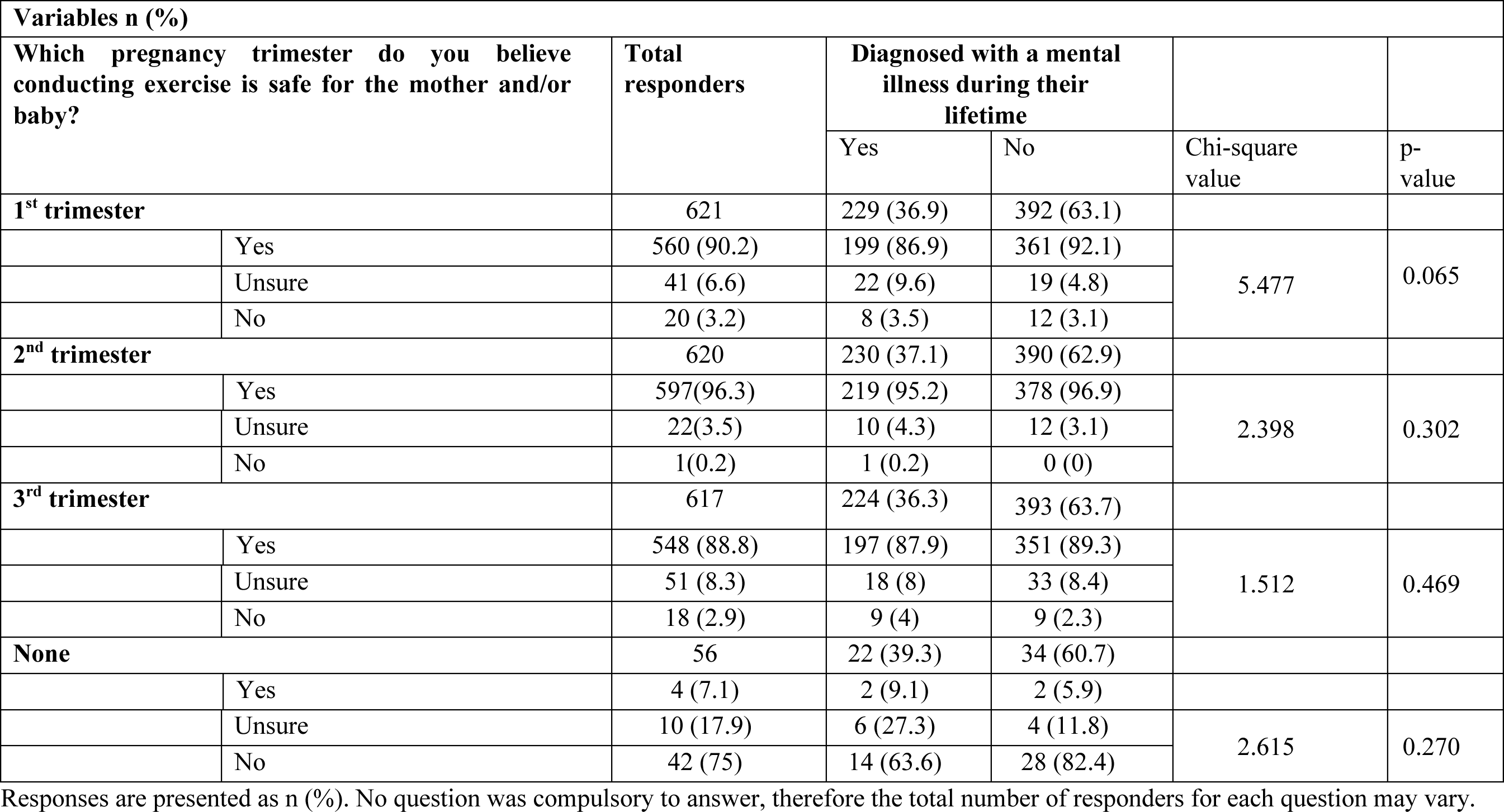
Perspectives about safety of conducting exercise during each pregnancy trimesters.

**Table S3:**
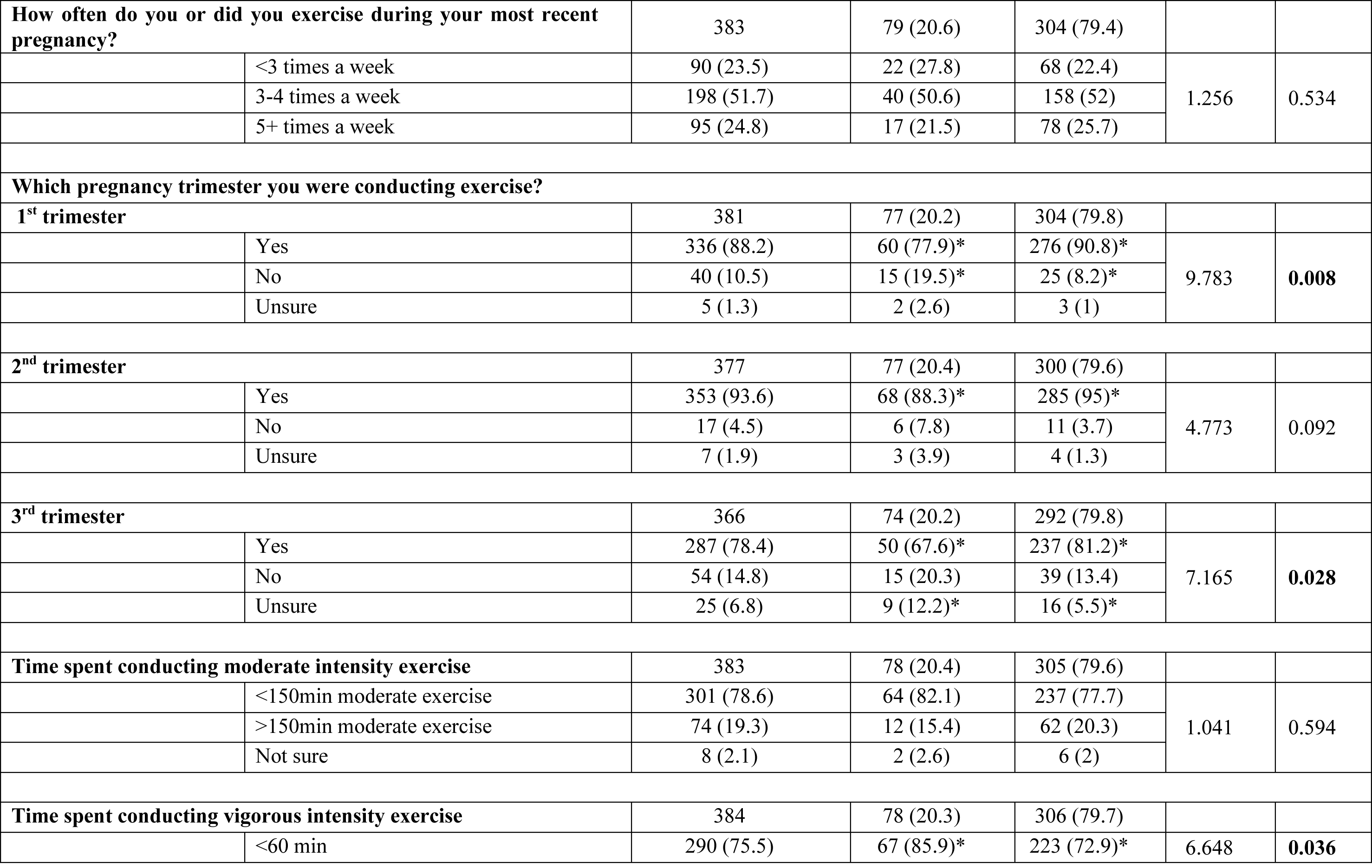
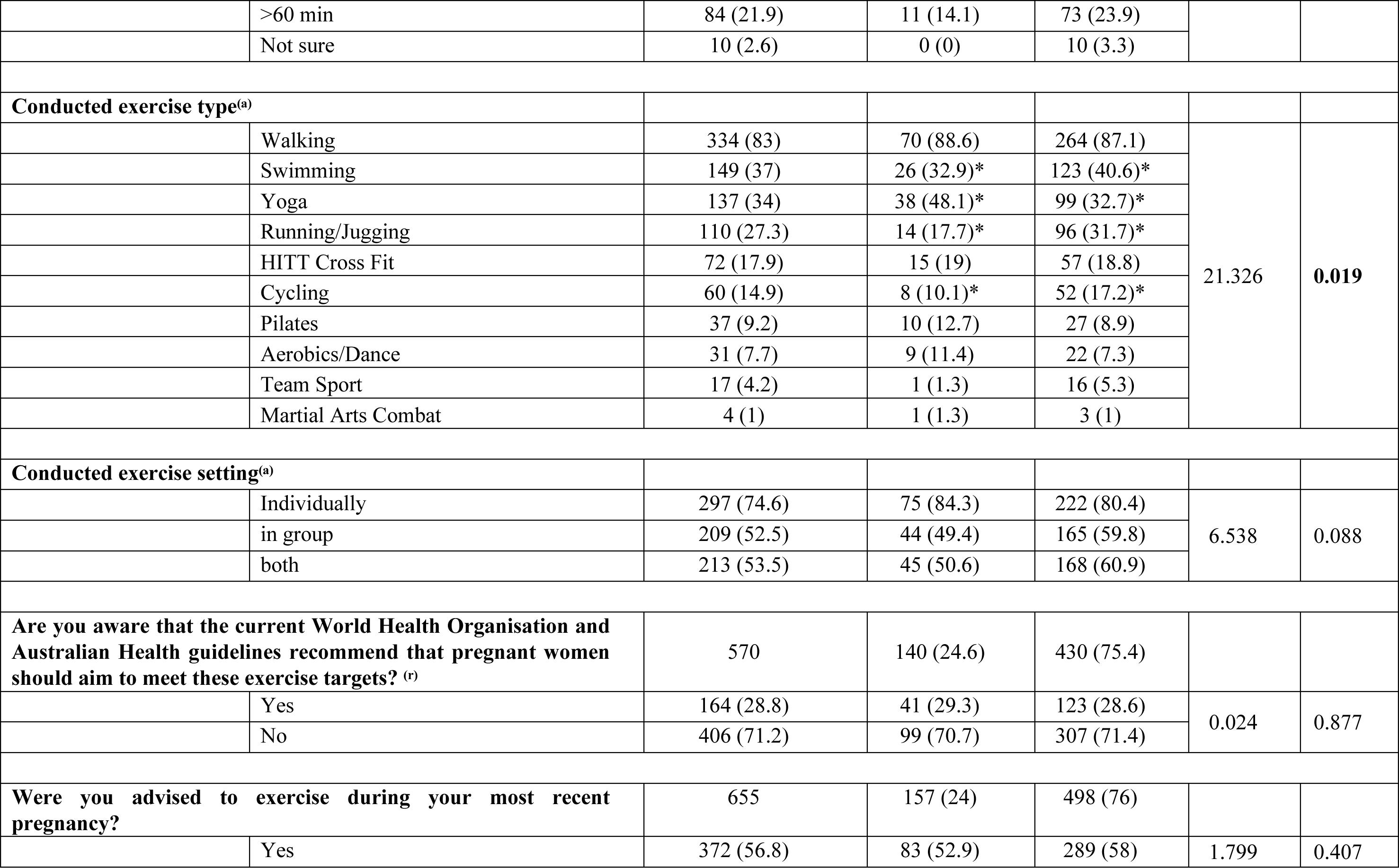
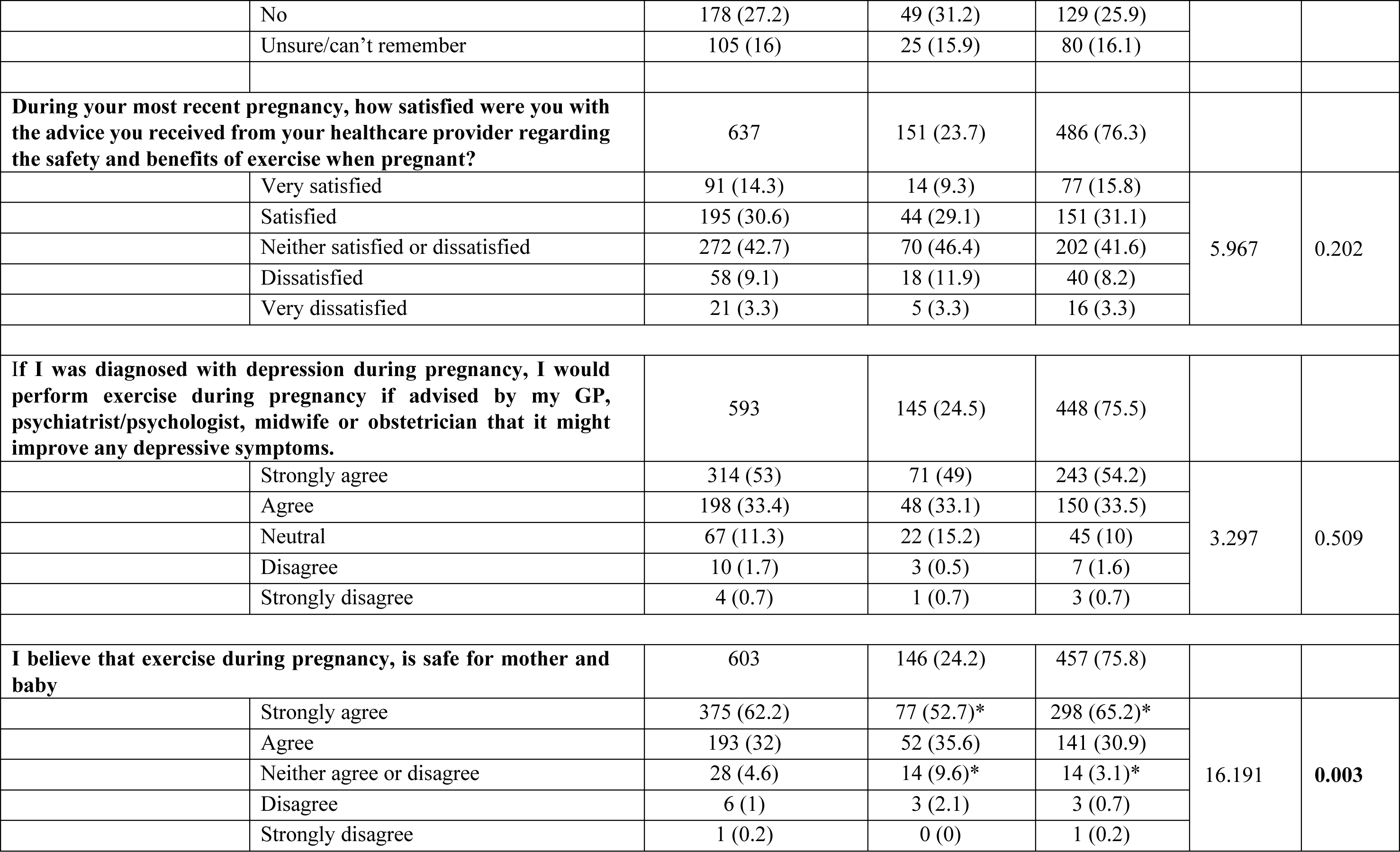
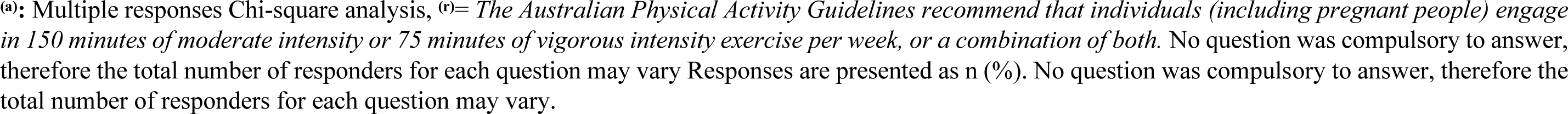
Exercise experience and advice received about exercise during the most recent pregnancy, in those that did and did not experience mental illness during pregnancy.

